# A Cognitive Development Chart for School-age Children and Adolescents

**DOI:** 10.1101/19012963

**Authors:** André R. Simioni, Daniel S. Pine, João R. Sato, Pedro M. Pan, Rochele Paz Fonseca, Julia Schafer, Euripedes C. Miguel, Jair J. Mari, Ary Gadelha, Rodrigo A. Bressan, Luis A. Rohde, Giovanni A. Salum

## Abstract

**Objective:** To evaluate the reliability and predictive utility of a time-efficient cognitive development chart that seeks to identify children and adolescents with high-risk for multiple outcomes such as mental health problems, substance use, and educational difficulties.

**Method:** We analyzed data from the Brazilian High-Risk Cohort for Psychiatric Disorders (HRC), a longitudinal school-based study conducted from 2010-2011 to 2013-2014. Participants were 2,239 children and adolescents, 6 to 17 years of age, who completed the cognitive assessment at baseline. The task used to track cognitive development was the Two Choice Reaction Time task (<3 minutes of duration, computer-based), which assesses the accuracy and speed of perceptual decision-making. Mental health, substance use, and educational outcomes were assessed by validated standardized methods. Key variables were measured at baseline and 3-year follow-up. The predictive utility was assessed using static (deviations from the age-expected performance at baseline) and dynamic (deviations from the age-expected change in performance over time) indicators.

**Results:** The reliability of the task parameter was high (intra-class correlation coefficient = 0.8). Static indicators of cognitive development significantly predicted concurrent mental, intellectual and educational difficulties, as well as incident and persistent educational difficulties and substance use in the 3-year follow-up. Dynamic indicators predicted persistent mental health problems.

**Conclusion:** Primary-care and mental health professionals need a time-efficient tool for tracking deviations from age-expected cognitive development, which predicts multiple unwanted outcomes at the same time. If replicated, future results could support the generation of tools for tracking risk for mental health, substance use, and educational difficulties.

## INTRODUCTION

Tracking physical health through normative growth charts is a routine component of pediatric care and have proved valuable in the early identification of health problems. Tools such as the World Health Organization (WHO) Growth Chart standards quantify the adequacy of growth under optimum conditions and exposure to few if any risk factors.^1^ It became worldwide adopted due to it’s inexpensive, safe, low-training-requirement properties and easy-to-interpret graphical interface.

However, there are no universally implementable tools for routinely tracking cognitive development. The available ones have problems.^2–7^ They assess participants of a limited age range, are costly, culturally bound, complex in administration and scoring, or possess limited data on predictive validity. Given primary-care and mental health professionals are interested in overall healthy development and not on preventing specific disorders/situations, there is a great need for time-efficient tools that can predict multiple outcomes at the same time. A recent meta-analysis found the average primary-care physician consultation to range in duration from 48 seconds in Bangladesh to 22.5 minutes in Sweden; 18 countries reported mean consultation lengths of 5 minutes or less.^8^ These facts underscore the need for brief screening tools with predictive validity.

Cognitive abilities predict many real-world outcomes, including education, occupation, and income^9^, social functioning^10^, physical and mental health outcomes^11^. Processing speed may represent one of the best predictors of overall cognitive abilities^12^. It has been longitudinaly linked to the development of resoning^13^ and it is much related to cognitive efficiency^14^. It can be assessed through a choice reaction-time task for simple decisions (e.g. the direction towards which an arrow points to), generating measures of response times and accuracy. This 3-minutes task has been used in psychopathology research^15^ to estimate cognitive and neural processes, making it suitable for routine screening in primary care and mental health settings. The current study describes the use of this task.

As far as we know no studies have showed mental health, substance use and educational outcomes predicted altogether using a unique screening 3-minutes task mapping cognitive efficiency. This study has three objectives: (1) to test the short-term reliability of a Cognitive Development Chart constructed to investigate processing efficiency in school-age children and adolescents; (2) to investigate if deviations from age-expected performance derived from the chart (static prediction) are associated with concurrent and future mental health problems, substance use and educational difficulties, and (3) to investigate if deviations from age-expected change in performance from baseline to follow-up (rate of change, dynamic prediction) derived from the chart are associated with the same outcomes.

## METHODS

### Study design and participants

The Brazilian High-Risk Cohort for Psychiatric Disorders is a large longitudinal school-based study investigating typical and atypical developmental trajectories of psychopathology and cognition. From a screening phase including 9,937 biological parents of children aged 6 to 15 years registered at 57 schools in two Brazilian cities (Porto Alegre and São Paulo) interviewed with the Family History Screen^16^, we selected two sets of subjects to be further evaluated in the baseline assessment (2010-2011). One recruited using a random selection (representative of the community, n=958) and the other using a high-risk selection procedure (subjects at increased risk for mental disorders based on family members’ and children’s symptoms, n=1,553) totalizing 2,511 subjects. We also conducted a 3-year follow-up with an 80% retention rate (wave 1, 2013-2014). For the present analysis, the sample was comprised of 2,239 subjects who completed the cognitive assessment at baseline. The Ethics Committee of the Universidade de São Paulo and Universidade Federal do Rio Grande do Sul approved the study. More details have been published elsewhere.^17^

### Measures

#### Cognitive Assessment

*Two-choice Reaction Time task (2C-RT)*, was the main task used to track cognitive development. The task is significantly associated with age (Figure S1) and it is the shortest task used in our cognitive battery. It measures the speed/accuracy to discriminate the direction (left/right) to which an arrow is pointing at, on the computer screen (100 trial with equal probabilities).^18^ Data on mean reaction time and accuracy were used as input for an EZ-diffusion model function (Figure 1), from here.^19^ This model assumes that a decision is made through the accumulation of noisy evidence about a stimulus over time until one of the response criteria is reached. The most relevant parameter from the model is the drift rate (*v)*, which reflects the processing efficiency to discriminate a stimulus (high number means better processing efficiency). This measure has been associated with several forms of psychopathology.^15^ Therefore, it was the chosen parameter for developing our chart.

**Figure 1:**
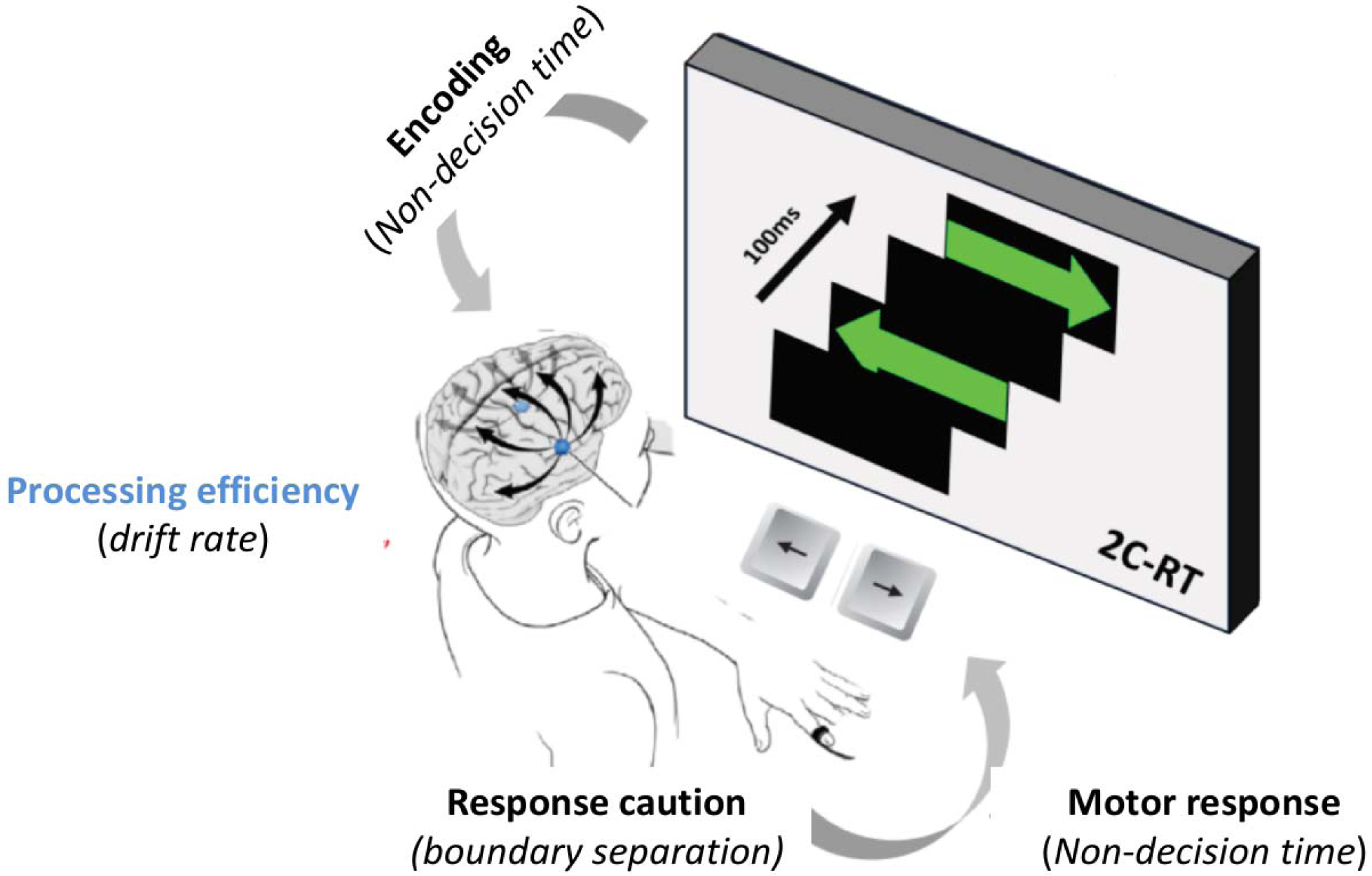
EZ-diffusional model components. Note: This model considers speed and accuracy simultaneously. *Non-decision time* represents the encoding and motor response time; *boundary separation*, the amount of information needed before making a decision (response caution style); and *drift rate*, the average rate of evidence accumulation within a trial.

Additionally to the static assessment, the rate of change of processing efficiency was also calculated - parameter from follow-up minus baseline divided by the number of years between. Processing efficiency is expressed in processing efficiency units (PU), which is a standardized score using the typically developing children as the reference group.

*Conflict Control task (CCT)* is very similar to 2C-RT, but it includes a second inhibitory component.^18^ This task included 75 congruent trials when participants had to indicate the actual green arrows’ directions, in addition to 25 incongruent trials when participants had to respond in the opposite direction of red arrows. The EZ-diffusion model processing efficiency component of congruent trials of CCT was used to assess the 2C-RT short term test-retest reliability. In half of the children, the task was administered before the 2C-RT and half of them after the 2C-RT.

#### Outcomes

Mental health and educational outcomes were assessed by validated standardized interviews and tests performed by trained psychologists and speech therapists.

Mental Health Assessment

- *Child psychiatric diagnoses* were assessed using the Brazilian version of the Development and Well-being Assessment^20^, a structured interview with items based on the DSM-IV-TR. At baseline, only the primary caregiver answered the instrument; in wave 1, the parental interview was complemented with a child semi-structured interview on internalizing disorders. Both responses generate an initial computerized diagnosis that can be confirmed, refuted or altered by trained child psychiatrists who ascertain final diagnoses. A second child psychiatrist rated a total of 200 interviews from the study, which resulted in a high inter-rater agreement (k-value=0.80, expected agreement=54.6; rater agreement=90.95).^17^ For this analysis, we used only any mental disorder category (Table S1).
- *Underdeveloped intellectual abilities* were defined as an intelligence quotient below 80, estimated according to the Tellegen and Briggs method^21^ and vocabulary and block design subtests of the Weschler Intelligence Scale for Children, third edition – the only existing Brazilian version till 2010.^22^ This measure was only available at the baseline assessment.

Substance Use

- *Current substance-use:* questions about the past 12 months’ use of alcohol, tobacco, shoe glue, solvents, crack, cocaine, marijuana, ecstasy, acids, amphetamines, steroids or other recreational drugs were presented to the child at wave 1. This variable was defined as “present” if the participant answered positively to any substance-use over the past 12 months.

Educational outcomes

- *Academic difficulties* were assessed by certified speech therapists with the reading (recognition of 70 isolated words) and writing (34 isolated words in dictation) subtests of the Brazilian School Performance Test.^23^ Reading time was also recorded. On wave 1, the test was abbreviated (61 words from reading and 13 from writing subtests) to only include highly informative items, selected using Item Response Theory.^23^ On both assessments, a unidimensional confirmatory factor analysis model (CFA) with each of these dimensions was built, and individuals’ standardized factor scores were saved regressing out the effects of age and gender. This score was then converted to centiles and defined as reading or writing difficulty if the subject’s centile was below 5% and as reading speed difficulty if the subject’s centile was above 95%.
- *School achievement* was assessed by the items about school performance of the Brazilian Portuguese version of the Child Behavior Checklist (CBCL) for ages 6 to 18^20^ completed by the caregiver. Participants were scored as failing, below average, average, and above average in the following academic subjects: Portuguese or literature, history or social studies, English or Spanish, mathematics, biology, sciences, geography, and computer studies. A composite score with these items was derived from saved standardized factor score from an unidimensional CFA described elsewhere.^24^ The factor score was converted to centiles and declared as school achievement difficulty if the subject’s centile was below 5%.
- *Difficulty in educational performance* reflects any difficulties in reading accuracy, reading speed, writing accuracy and school achievement previously described.
- *Reading comprehension* was assessed by asking the participant to read a school text of general knowledge without time limitation and answer orally to ten text comprehension questions.^25^ The trained evaluator rated accuracy. An unidimensional CFA model with these items was fitted and individuals’ standardized factor scores were saved regressing out the effects of age and gender. This score was then converted to centile and defined as reading comprehension difficulty if the subject’s centile was below 5%. This variable was assessed only at the follow-up.

#### Other Assessments

To generate the task parameters from the 2C-RT in typically developing children, we selected children not exposed to well-known risk factors for mental health problems and educational difficulties, which were assessed as described below.

- *Exposure to Child Abuse/Neglect* was evaluated by asking both, caregiver and children, the frequency of lifetime physical neglect and physical, emotional and sexual abuse. A CFA was built and the factor scores were used to determine the presence/absence of trauma exposure as described in detail elsewhere.^26^
- *Exposure to prenatal and perinatal risk factors* were evaluated asking the parental respondent at how many gestational weeks the child was born, the frequency at which the mother smoked during the pregnancy and if someone else smoked near her during that period.
- Socioeconomic status was evaluated according to the *Associação Brasileira de Empresas de Pesquisa* criteria from 2009. We merged classes A and B into a high status, C into a medium status, and D and E into a low socioeconomic status.
- *The current parental psychiatric diagnosis* was assessed by applying the Mini International Neuropsychiatric Interview^27^ to the respondent (biological mother in 92% of the cases). An ‘any mental disorder’ variable was created encompassing any anxiety, mood, substance use disorder, psychosis or ADHD.

### Data Analysis

#### Reliability analysis

To estimate test-retest levels, we compared the processing efficiency diffusion model-components from the 2C-RT and the congruent trials of the CCT, which use identical stimuli. The Bland-Altman method^28^ was used for analyzing the agreement between the measures by constructing limits of agreement which are calculated using the mean and the standard deviation (SD) of the differences between the two measures. It is recommended that 95% of the data points should lie within ± 1.96 SD of the mean difference. The lower the range between these two limits the better the agreement is. Additionally, the intra-class correlation coefficient (ICC) was estimated using a two-way mixed effect (agreement, average-measures) from the *irr* package version 0.84.^29^ ICC ranges from 0 (low) to 1 (high) indicating test-retest similarity.

#### Deriving the Cognitive Development Chart

Participants were split into a typically developing sample (n=196) and a validation sample (n=2043). The typically developing sample was composed of randomly-selected participants from the community, pertained to medium to high socioeconomic status, born after 37 gestational weeks, without exposure to abuse, neglect, gestational smoke, and with no personal, or parental history of a mental disorder. We aimed to assess cognitive development in the absence of known risk factors for psychopathology, a strategy also used in WHO growth charts.^1^

Age-standardized curves of cognitive development were generated at baseline and follow-up using age as an independent variable and processing efficiency as a dependent variable in a Generalized Additive Model (GAM, from *gamlss* R package, version 5.1-2).^30^ Next, we tested linear, quadratic and cubic terms and selected the best-fit ones according to the lowest scores of Akaike Information Criterion (AIC) and Schwartz Bayesian Criterion (SBC). The best-fitting GAM to the training sample was the linear model (SBC = −1872, AIC = −1881) compared to quadratic (SBC = −1867, AIC = −1880) and cubic (SBC = −1865, AIC = −1879) terms. Lastly, we extracted the predicted values in the validation sample and standardized it using the parameters from the typically developing sample.

#### Testing associations with mental health and educational outcomes

Deviations based on the standardized centiles were used as independent variables to predict concurrent and 3-year outcomes. Logistic regression and signal detection analyses were performed, investigating the saved predicted probabilities for each centile of the processing efficiency variable. All analysis were conducted in R version 3.5.1, including *cutpointr* package version 0.7.4.^31^

## RESULTS

Overall, at baseline, our sample was equally distributed according to sex. It had a mean age of 10 years and was derived from mid-level socioeconomic-status households. One-fourth of the participants were diagnosed with a mental disorder and one-seventh were classified as having difficulties in educational performance at baseline and one-third used any substances at follow-up (Table 1). As expected, processing efficiency increased with development (Figure S2, panel A).

**Table 1:** Descriptive table.

|  | Typically Developing Sample |  | Validation Sample |  | Total |  |
| --- | --- | --- | --- | --- | --- | --- |
|  |  | N |  | N |  | N |
| **Sociodemographic** |  |  |  |  |  |  |
| Age |  | 196 |  | 2043 |  | 2239 |
| - Mean (SD) | 10.32 (2.00) |  | 10.41 (1.94) |  | 10.41 (1.94) |  |
| - Range | 6.80 - 14.53 |  | 5.88 - 14.43 |  | 5.88 - 14.53 |  |
| Male | 107 (54.6%) | 196 | 1114 (54.5%) | 2043 | 1221 (54.5%) | 2239 |
| **SES** |  |  |  |  |  |  |
| - High | 105 (53.6%) |  | 790 (38.7%) |  | 895 (40.0%) |  |
| - Medium | 91 (46.4%) |  | 1181 (57.8%) |  | 1272 (56.8%) |  |
| - Low | 0 (0.0%) |  | 72 (3.5%) |  | 72 (3.2%) |  |
| **Baseline Assessment** |  |  |  |  |  |  |
| **Mental Disorders** |  |  |  |  |  |  |
| Any Mental Disorder | 0 (0.0%) | 196 | 606 (29.7%) | 2043 | 606 (27.1%) | 2239 |
| Underdeveloped Intellectual Abilities | 12 (6.2%) | 195 | 164 (8.7%) | 1888 | 176 (8.4%) | 2083 |
| **Academic Performance** |  |  |  |  |  |  |
| Any Difficulty in Educational Performance | 26 (13.3%) | 196 | 296 (14.5%) | 2043 | 322 (14.4%) | 2239 |
| **Follow-up Assessment** |  |  |  |  |  |  |
| Current Substance Use | 41 (29.9%) | 137 | 417 (29.2%) | 1427 | 458 (29.3%) | 1564 |
| Reading Comprehension Difficulties | 8 (5.6%) | 143 | 193 (12.3%) | 1567 | 201 (11.8%) | 1710 |
| **Incident** |  |  |  |  |  |  |
| Any Mental Disorder | 21 (14.1%) | 149 | 205 (17.6%) | 1164 | 226 (17.2%) | 1313 |
| Any Difficulty in Educational Performance | 8 (4.7%) | 170 | 237 (13.6%) | 1747 | 245 (12.8%) | 1917 |
| **Persistent** |  |  |  |  |  |  |
| Any Mental Disorder | 0 (-) | 0 | 204 (40.3%) | 506 | 204 (40.3%) | 506 |
| Any Difficulty in Educational Performance | 16 (61.5%) | 26 | 208 (70.3%) | 296 | 224 (69.6%) | 322 |

Notes: The training sample was composed of randomly-selected participants from the community, born after 37 gestational weeks, without exposure to abuse, neglect, gestational smoke, with no personal or parental history of a mental disorder and that pertained to medium to high socioeconomic status. Difficulties are defined as being below the 5th centile in each task. Reading Comprehension Test and Recent Substance Use were only evaluated at follow-up. An expanded version of this table, with sub-diagnosis and educational subcategories, is available in Table S1. Abbreviations: SES, socioeconomic status; SD, standard deviation.

### Test-retest reliability

The ICC indicates moderate-to-good short-term test-retest reliability (0.80, IC95% 0.70-0.86, Figure 2, panel B). Similarly, the mean difference (-0.0012), the narrow limits of agreement of the processing efficiency component on the Bland-Altman plot and the low rate of change per-year (mostly around ± 0.5 z-scores) suggest very-good agreement (Figure S3-4).

**Figure 2:**
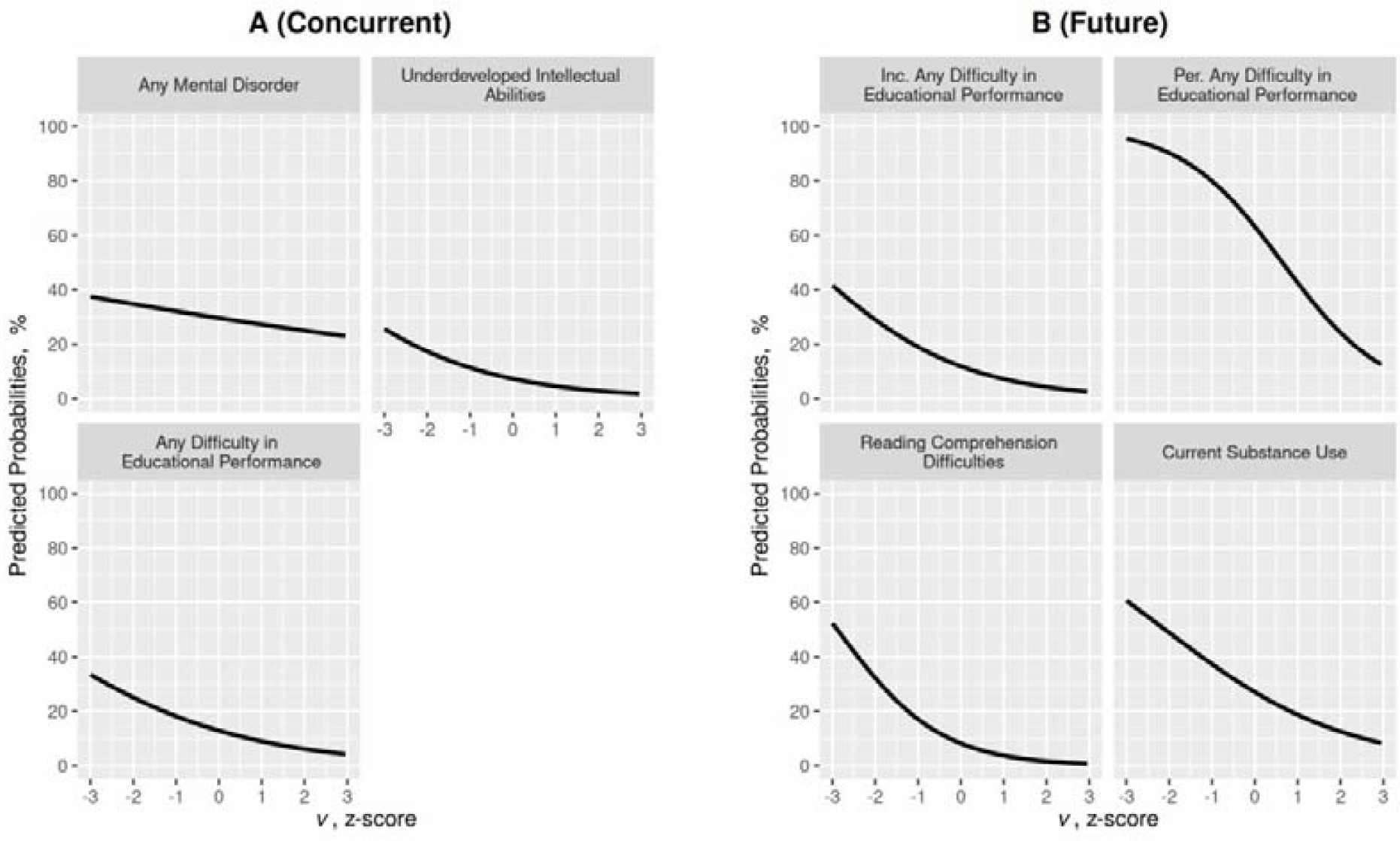
Static prediction of concurrent and future mental health, substance use and educational outcomes. Notes: Predicted probabilities of significant concurrent (panel A) and future (panel B) mental health problems and educational difficulties according to performance on the processing efficiency (*v*, static prediction).

### Predicting probabilities of negative outcomes using the Cognitive Development Chart

#### Static predictions

##### Concurrent associations

At baseline, deviations from age-expected performance were associated with any mental disorder (Table S2, OR=0.89, 95%CI 0.81-0.97, p=0.008), underdeveloped intellectual abilities (OR=0.61, 95%CI 0.53-0.70, p<0.001) and educational-performance difficulties (OR=0.66, 95%CI 0.59-0.75; p<0.001). Participants with −2 and +2 z-scores on the curve had the following predicted probabilities: any mental disorder, 35% and 25%; underdeveloped intellectual abilities, 19% and 1%; and educational-performance difficulties, 25% and 5% (Figure 2, Panel A).

##### Future outcomes

Deviations from age-expected performance at baseline significantly predicted incident difficulties in educational performance (Table S3, OR=0.57, 95%CI 0.50-0.65, p<0.001), difficulties in reading comprehension (OR=0.43, 95%CI 0.37-0.50, p<0.001) and substance-use (OR=0.62, 95%CI 0.56-0.69, p<0.001). Besides, performance on the task also predicted the persistence of educational difficulties (OR=0.43, 95%CI 0.32-0.57, p<0.001). Participants with −2 and +2 z-scores on the curve had the following predicted probabilities: incident difficulties in educational performance, 30% and 5%; persistent educational difficulties, 90% and 25%; difficulties in reading comprehension, 30% and 1% and substance-use 50% and 10% (Figure 2, Panel B).

##### Dynamic predictions

Deviations from the age-expected change in task performance over time significantly predicted the persistence of mental disorders (OR=0.5, 95%CI 0.26-0.94, p=0.03). Participants with a change of − 1 and +1 z-scores per-year on the curve had 61% and 28% of predicted probabilities for persistent mental disorder (Figure 3). The signal detection analysis of these results can be found at Table S4-7.

**Figure 3:**
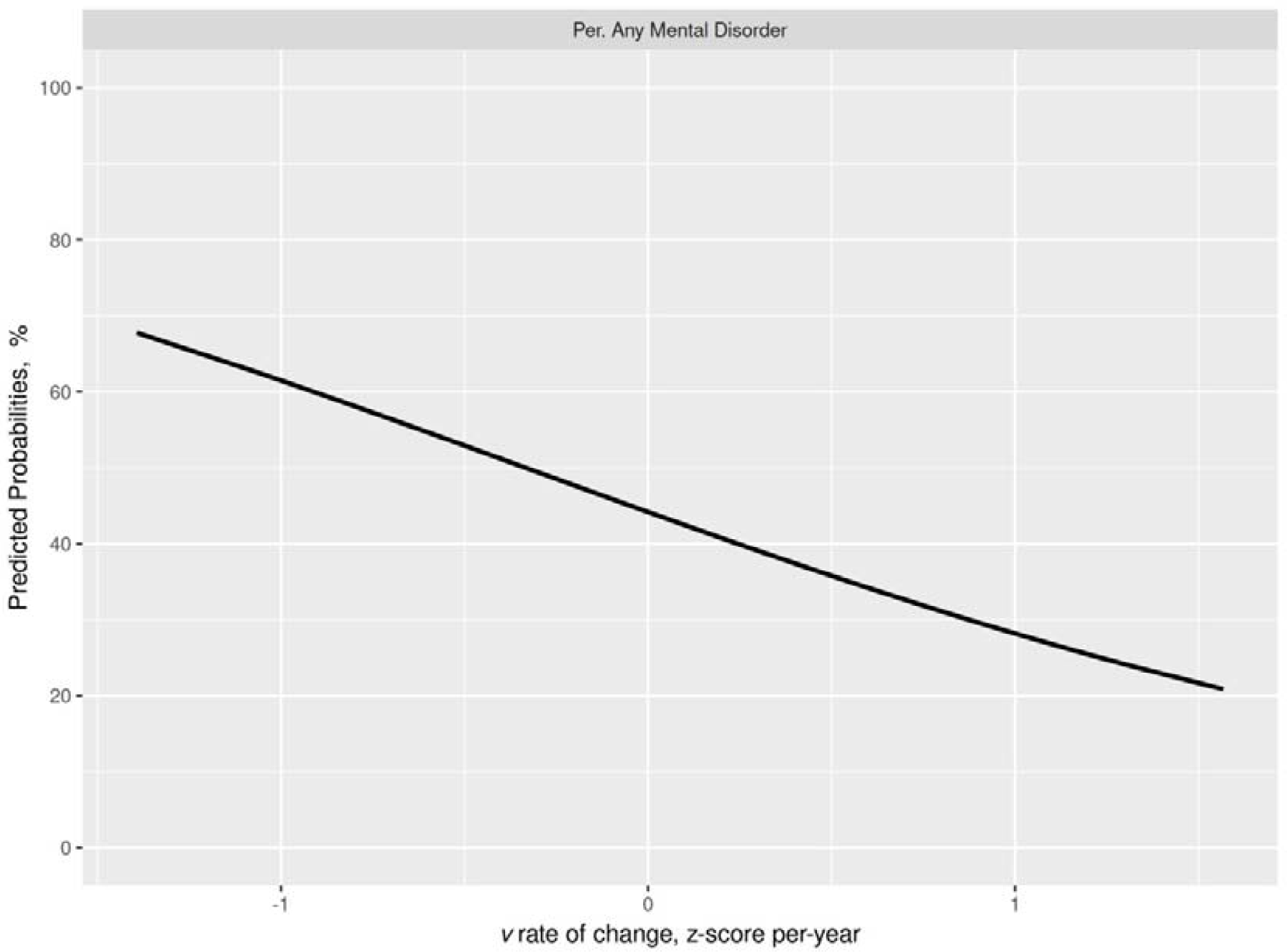
Dynamic prediction of future mental health problems. Note: Predicted probabilities of significant future mental health problems according to rate of change over time (z-score per-year) of the processing efficiency (*v*) from the baseline to the follow-up assessment (dynamic prediction)

**Figure 4:**
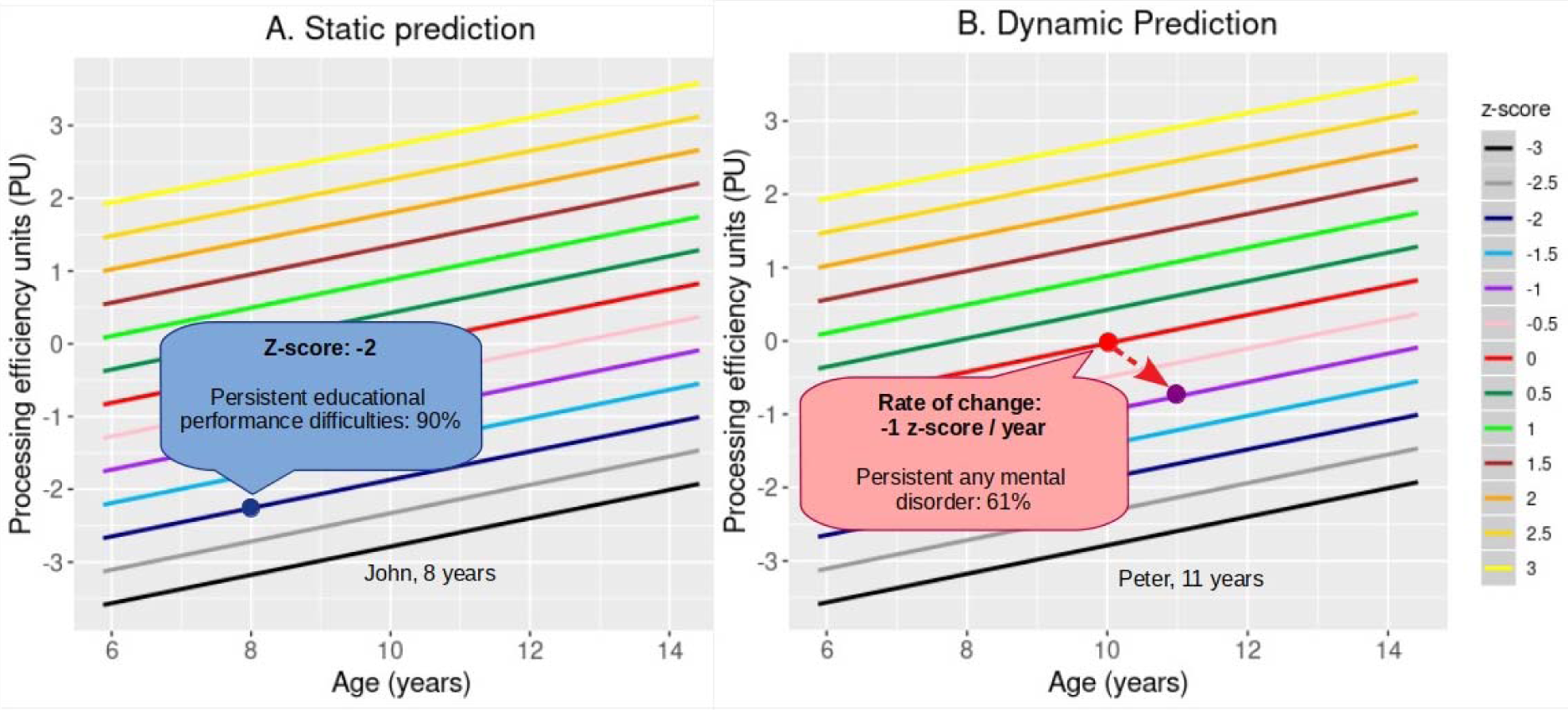
Cognitive development chart examples. Notes: PU against age, with colors representing z-scores. The child PU is plotted against his/her age to locate the z-score position. With this position is possible to estimate the probability of concurrent or future mental health, substance-use or educational outcomes by checking the predicted probabilities of these outcomes (Figures 2-3).

## DISCUSSION

This study describes the creation and validation of a screening tool, the Cognitive Development Chart, for use by primary-care and mental health professionals seeking to identify children and adolescents with ongoing or risk for future mental health, substance use, and educational difficulties. The chart demonstrated acceptable test-retest reliability and predictive utility for tracking many important aspects of children’s development.

Current methods for tracking mental health and cognitive risk utilize a piecemeal approach. In the mental health field, most work investigating screening tools focuses on symptom scales, such the Pediatric Symptom Checklist, CBCL, and the Strengths and Difficulties Questionnaire.^3–5,7,20^ Such scales effectively identify risk for mental disorders. However, they fail to quantify the continuous age-related changes in developmental competencies and often rely on parental reports, which can reduce accuracy through parental psychopathology or confirmation, and social desirability biases. Moreover, the tools require some time to complete, and focus primarily on problems in the mental health domain.

In the educational field, most tools assess literacy directly through measures, such as the Dynamic Indicators of Basic Early Literacy Skills (DIBELS)^32^, and the Computer-based Phonologic Awareness Screening and Monitoring Assessment (Com-PASMA)^6^. While useful, these tasks have limitations. For example, the DIBELS can have floor effects that compromise predictive ability^32^, while the Com-PASMA only targets preschoolers. Recently, brain function measures (neuroimage and cognitive tasks) had been studied through a growth chart analysis providing a novel approach for constructing biomarkers for mental health relevant conditions.^33–35^ However, these and other cognitive tasks can be time-consuming (with at least one-hour testing) with complex administration requirements.

The Cognitive Development Chart described in the current report may provide a cost and time-effective, easy-to-acquire, and reliable cognitive measure that could complement the growth monitoring routine. Many primary-care and mental health professionals fail to use currently-available mental-health screening tools.^36^ The availability of time-efficient measures could address the need for scalable tools^37^ that support the broad dissemination of effective practices. Unlike currently-available ones, this new measure generates predictive information on risk for multiple adverse outcomes, including mental disorders, substance use and difficulties in educational performance. Also, the results are in line with other studies associating processing speed with psychopathology^38^ and academic difficulties.^13,14^ Finally, given the widespread adoption of the WHO standard curves, this strategy might be particularly interesting and scalable to build informed health care systems.

This study has some limitations. *First*, longitudinal trajectories were estimated using two-time points with an average of 3-years between them and with multiple subjects’ ages. It is unclear whether measures over shorter periods would have similar predictive abilities. *Second*, the analysis was limited to one cohort and still needs to be determined whether results would replicate in less controlled settings. *Third*, reliability analysis were performed using distinct tasks in a short period. However, using distinct tasks constitutes a conservative bias since it is unlikely reliability would be lower using the same task. Nevertheless, this study has important strengths. *First*, we took advantage of a very simple tool, with a low interviewer effect, which was able to predict multiple outcomes, such as mental health problems, substance use and educational difficulties in a large sample of children. *Second*, a more dynamic measure of development (rate of change) also predicted negative mental health outcomes, which support this measure as an overall tracking tool for a variety of problems in school-age children.

## CONCLUSION

We described an efficient screening method, which efficiently identifies children with concurrent mental health problems and high risk for future substance use and educational problems. If replicated, these results could inform attempts to track mental health and cognitive development using a scalable strategy that identifies children likely to benefit from selective prevention efforts.

## Data Availability

Data is available under request.

## Abbreviations

2C-RT: Two Choice Reaction Time Task
AIC: Akaike Information Criterion
CBCL: Child Behavior Checklist
CCT: Conflict Control task
CFA: Confirmatory Factor Analysis Model
GAM: Generalized Additive Model
ICC: Intra-class Correlation Coefficient
SBC: Schwartz Bayesian Criterion
SD: Standard Deviation
PU: processing efficiency units
WHO: World Health Organization

## Notes

**Financial support** Financial Disclosure: Prof Pan receives a post-doctoral scholarship from FAPESP. Prof Salum received research grants from national funding agencies: FAPERGS, CAPES, and CNPq. Prof Miguel and Bressan received research grants from national funding agencies: FAPESP, CAPES, CNPq. Prof Bressan also received research grants from Janssen Cilag, Novartis, Roche in the last 5 years. Prof Bressan has participated in speaker bureaus for Ache, Janssen, Lundbeck, and Novartis and has been a consultant for Janssen, Novartis, and Roche. Prof Rohde was on speakers’ bureau and/or acted as a consultant for Eli-Lilly, Janssen-Cilag, Medice, Novartis, receives authorship royalties from Oxford Press and ArtMed, and has received unrestricted educational and research support from Eli-Lilly, Janssen-Cilag, Shire, and Novartis.

Funding Information: This work is supported by the National Institute of Developmental Psychiatry, a science and technology institute funded by Conselho Nacional de Desenvolvimento Científico e Tecnológico (CNPq; National Council for Scientific and Technological Development; grant number 573974/2008-0 and 465550/2014-2) and Fundação de Amparo à Pesquisa do Estado de São Paulo (FAPESP; Research Support Foundation of the State of São Paulo; grant number 2008/57896-8 and 2014/50917-0). The funders did not interfere in data collection, analysis, design, or interpretation of the study.

**Conflict of interests:** The authors have no conflicts of interest relevant to this article to disclose.

### Competing Interest Statement

The authors have declared no competing interest.

### Funding Statement

Financial Disclosure: Prof Pan receives a post-doctoral scholarship from FAPESP. Prof Salum received research grants from national funding agencies: FAPERGS, CAPES, and CNPq. Prof Miguel and Bressan received research grants from national funding agencies: FAPESP, CAPES, CNPq. Prof Bressan also received research grants from Janssen Cilag, Novartis, Roche in the last 5 years. Prof Bressan has participated in speaker bureaus for Ache, Janssen, Lundbeck, and Novartis and has been a consultant for Janssen, Novartis, and Roche. Prof Rohde was on speakers' bureau and/or acted as a consultant for Eli-Lilly, Janssen-Cilag, Medice, Novartis, receives authorship royalties from Oxford Press and ArtMed, and has received unrestricted educational and research support from Eli-Lilly, Janssen-Cilag, Shire, and Novartis.
Funding Information: This work is supported by the National Institute of Developmental Psychiatry, a science and technology institute funded by Conselho Nacional de Desenvolvimento Científico e Tecnológico (CNPq; National Council for Scientific and Technological Development; grant number 573974/2008-0 and 465550/2014-2) and Fundação de Amparo à Pesquisa do Estado de São Paulo (FAPESP; Research Support Foundation of the State of São Paulo; grant number 2008/57896-8 and 2014/50917-0). The funders did not interfere in data collection, analysis, design, or interpretation of the study.

